# Phenotypic grouping of Catheter-Associated *Escherichia coli* from COVID-19 isolation wards using Hierarchical clustering in Surabaya, Indonesia

**DOI:** 10.1101/2023.07.26.23293190

**Authors:** Daniel Edbert, Ni Made Mertaniasih, Pepy Dwi Endraswari

## Abstract

**Introduction:** Moderate to critical COVID-19 patients may be indicated for urinary catheter use due to the risk of immobility and ventilator or oxygen use. In intensive care units, 18-81.7% of all patients use a urinary catheter. Almost all patients with urinary catheters suffered from bacteriuria within 30 catheter-days. Hospital-associated isolate tracing is mainly performed using complex molecular tests that are not vastly available. This study aims to trace catheter-associated urinary tract infection (CAUTI) isolates using a common hierarchical clustering method that is vastly available

**Methods:** This is a descriptive study presenting a collection of *Escherichia coli* culture data performed by dr. Soetomo Public Hospital microbiology laboratory from March, 26^th^ 2020 to March, 31^st^ 2021. Hierarchical clustering was performed using statistical software using Ward’s clustering method.

**Results:** There are 36 *E*.*coli* associated with CAUTI. Isolate biochemistry profile and minimum inhibitory concentrations profiles were clustered into 3 clades for each profile. A total of 9 cluster combinations were found. Cluster-ID 1 was melibiose fermenters, Cluster ID 2 was a non-Arginine utilizer, and Cluster ID-3 was an Arginine utilizer. Cluster MIC A consists of third-generation Cephalosporin resistant isolates, Cluster MIC C was multi-susceptible isolates. The Chi-square test between cluster ID and MIC showed no significant differences between the number of isolates per group (X^2^, p = .430, CI = 95%).

**Conclusion:** CAUTI associated *E*.*coli* is divided into 9 clusters. This indicates no cluster dominates the isolates, thus CAUTI is not caused by hospital transmission but by normal flora carried by the admitted patient.

## Introduction

Hospitalized COVID-19 patients need a thorough diagnostic approach, especially in the management of critical and severe COVID-19 patients that can mimic systemic bacterial infection.^1^ Bacterial and fungal co-infections are contributors to high mortality in viral pandemics.^2^ Although the COVID-19 death toll is high,^3^ co-infections and hospital-associated infections are low in overall proportion. Ventilator-associated pneumonia is the highest hospital-associated infection in COVID-19 patients.^4-8^ Ventilator-associated pneumonia affected 27% of COVID-19 patients with pneumonia.^4^ Secondary co-infection can reach up to 50% in all severe patients.^9^ Reports showed that positive culture in COVID-19 patients is more likely to indicate culture contamination and device colonization related to device-associated infections.^7^

COVID-19 causes systemic inflammation that causes acute systemic reaction and activation of fibrosis.^10^ Death by COVID-19 is not directly caused by the viral pathogenesis, but also by the process of systemic inflammatory response and systemic fibrosis which impairs brain capability to receive hypoxia signals, and insufficient energy from aerobic metabolism is reduced.^10,11^ This permits more biofilm formation and bacterial invasion and increases the risk of secondary infection.

Limitation of patient care by healthcare personnel using layers of personal protective equipment contributes to the high number of specimen contamination and low patient care,^12^ especially in the early days of the COVID-19 pandemic. Blood culture contamination can reach as high as 65%.^7^

Severe and critical COVID-19 patients are susceptible to urinary tract infections due to urinary catheter use. Sedation and immobilization increase the use of urinary catheters. In non-COVID-19 intensive care, the use of urinary catheters is 81.7% of total admitted patients. The incidence of bacteriuria increases by 3-8% per catheter-days, and almost every patient contracts bacteriuria within 30 catheter-days.^13^ Most catheter-associated bacteriuria are asymptomatic and less than 5% develop into symptomatic bacteriuria. Catheter-associated urosepsis accounts for 15% of all nosocomial bacteremia and 10% of deaths associated with urosepsis.^14^

Biofilms are the common source of catheter-associated bacteriuria. Some efforts like antibiotic-impregnated catheters might reduce biofilm formation. The Planktonic phase of biofilm helps the bacteria to spread.^14,15^ this mobilization of biofilm-forming bacteria is important to identify and trace bacterial strains that spread. These strains can be traced using their genetic and phenotypic characteristics. Advanced methods of molecular tracing are available, but might not be readily available in limited-resource settings. Even though results of bacterial identification are often neglected, un-recorded, or only used in quality control measures it surely has clinical use. The probability of a strain showing certain characteristics in biochemistry tests reflects its genetic regulation as a response to exogenous exposure such as environmental stressors or past antimicrobial exposure.^16^ This can be used as the background to group these strains into clusters based on their phenotypic similarity.

This study aims to trace catheter-associated urinary tract infection (CAUTI) isolates using a common hierarchical clustering method that is easy and vastly available

## Methods

This is a descriptive study presenting a collection of urinary catheter-associated *Escherichia coli* culture and its patient-related data performed by the Microbiology laboratory of a COVID-19 referral hospital from March, 26th 2020 to March, 31st 2021 during the first wave of COVID-19 in Surabaya, Indonesia. Data was collected using a total consecutive sampling method from patients admitted to COVID-19 ICU. CAUTI cases were defined according to Symptomatic UTI (SUTI) 1a definition from NHSN patient safety component manual 2021.^17^

CAUTI cases in this research were defined by the following definitions: 1) Patient had an indwelling urinary catheter that had been in place for more than 2 consecutive days in an inpatient location on the date of event/date of sampling AND the catheter was either: Present for any portion of the calendar day on the date of event/date of sampling OR removed the day before the date of the event. 2) Patient has at least one of the following signs or symptoms: Fever (>38°C), suprapubic tenderness, costovertebral angle pain or tenderness, urinary urgency, urinary frequency, Dysuria. 3) Patient has a urine culture with no more than two species of organisms identified, at least one of which is a bacterial load of ≥10^5^ CFU/ml. All elements of the SUTI criterion must occur during the infection window period (3 days before and 3 days after onset).^17,18^

### Ethical statement

This research has been exempted after reviewing of dr. Soetomo public hospital Ethical committee letter number 0457/LOE/301.4.2/V/2021.

### Inclusion and exclusion criteria

Biochemistry and MIC data that were collected were the ones recorded using the BD Epicenter (Beckton-Dickinson) program and were performed using BD Phoenix. Any manual test results were excluded because of the risk of technical bias during the test performance. Clinical information that was collected was the ones recorded using the hospital information system. Missing data were excluded from the analysis

### Data Collection method and analysis

Biochemistry and MIC data were gathered using the BD Epicenter program and clinical information was collected using the hospital information system. Hierarchical cluster analyses were performed to analyze the similarity of isolates using biochemistry profile and Minimum Inhibitory Concentration (^2^Log_MIC_) from an automated identification system (BD Phoenix, Becton-Dickinson).^19,20^ was performed using SPSS version 27, in the classify tab, with Ward’s Method and Euclidian distance measurement. Binary data setting was used to analyze biochemistry profiles, Interval data setting was used to analyze Log _MIC_. After the cluster members were listed, the total isolates within the clusters were put in crosstabulation and Chi-square analyses were performed to analyze the differences of isolates between groups.

## Results

There are 77 *Escherichia coli* isolated from various specimens in COVID-19 isolation wards from March, 26^th^ 2020 to March, 31^st^ 2021 (Table 1). There were 36 *E*.*coli* isolated from urine specimens associated with CAUTI, another 15 were from urine specimens not associated with CAUTI, and the other 26 were from other anatomical sites. Mean catheter-days before specimen collection was 9.28 ± 1.37 days, with a median of 6 days. The longest catheter-days were 36 days and the shortest and the most occurring was 2 days.

**Table 1.** General characteristics of patients with CAUTI in COVID-19 isolation ward.

|  | N (%) |
| --- | --- |
| Sex |  |
| male | 10 (27.8) |
| female | 26 (72.2) |
| Source |  |
| Intensive care unit | 14 (38.9) |
| High care unit | 18 (50) |
| Low care unit | 4 (11.1) |
| Timing |  |
| March - June 2020 | 2 (5.5) |
| July - September 2020 | 10 (27.8) |
| October – December 2020 | 14 (38.9) |
| January – March 2021 | 10 (27.8) |
| Death associated with CAUTI | 13 (36.2) |

Isolate biochemistry profile and minimum inhibitory concentrations profiles were clustered into 3 clades for each profile. ID clusters were named with numbers, while MIC clusters were named with letters. A total of 9 cluster combinations were found similar distributions were noted in the number of cluster members (Table 2). After a retrospective analysis of the isolates, it can be identified that Cluster ID 1 was Melibiose fermenters and the most metabolically active cluster, Cluster ID 2 was non-Arginine utilizer and a less metabolically active cluster, and Cluster ID-3 was Arginine utilizer. Cluster MIC A consists of third-generation Cephalosporin resistant isolates, Cluster MIC C has multi-susceptible isolates (Figure 1). The Chi-square test between cluster ID and MIC showed no significant differences between the number of isolates per group (X^2^, p = .430, CI = 95%)

**Table 2.**
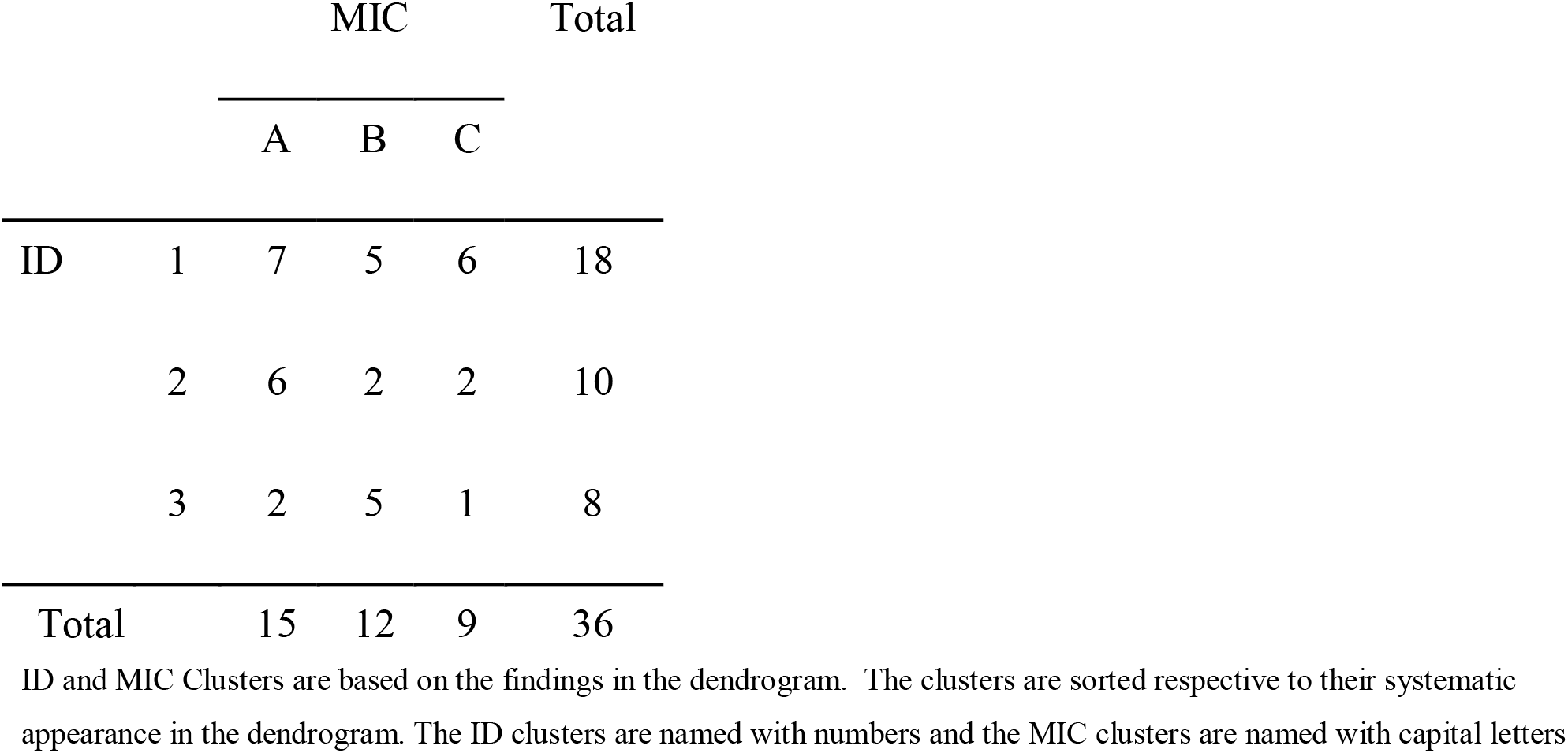
Cross-tabulation of ID and MIC clusters.

**Figure 1.**
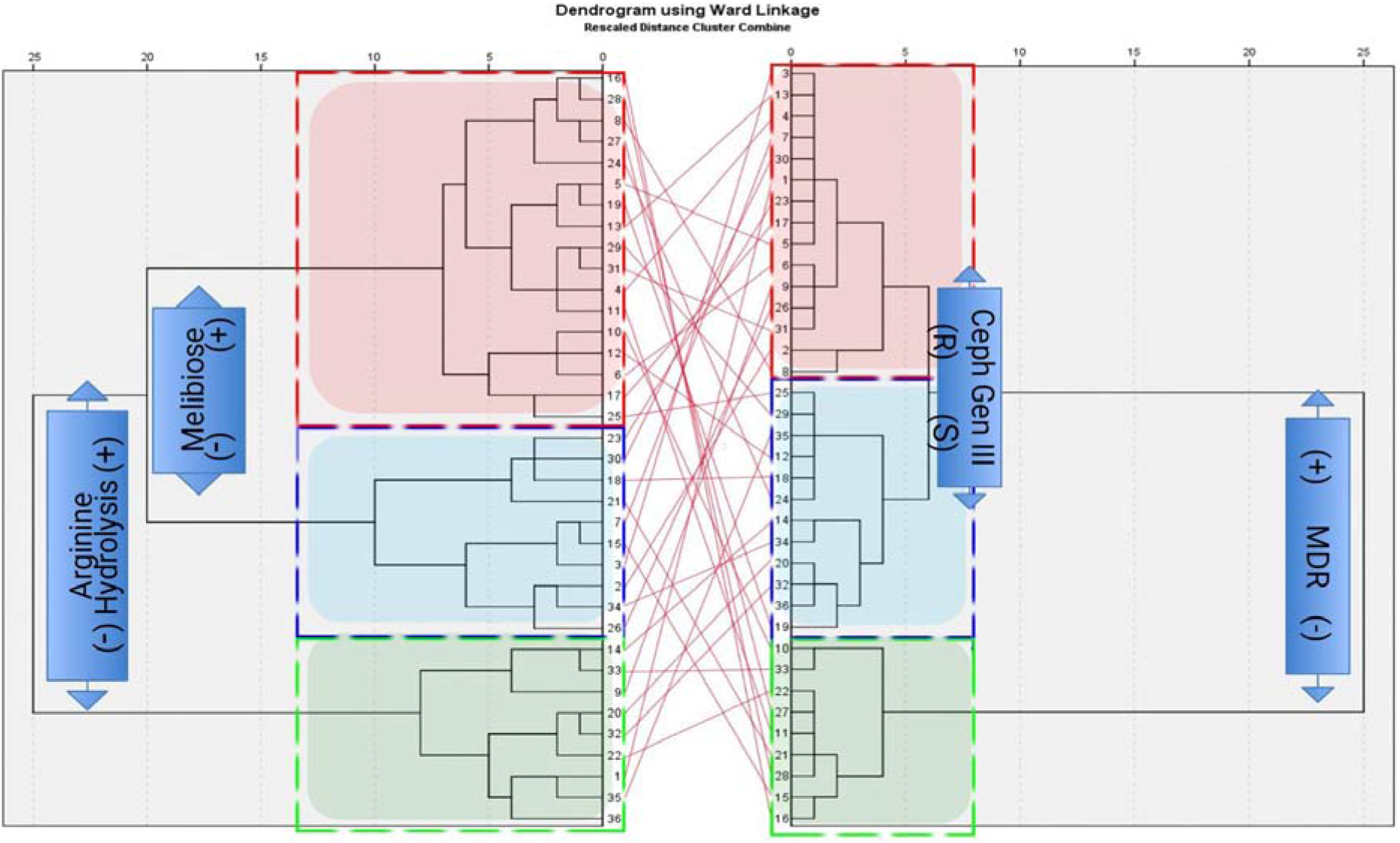
Left dendrogram is hierarchical clustering for biochemistry (ID clusters), and the right dendrogram is hierarchical clustering for ^2^Log_MIC_ (MIC clusters). There are 3 cluster clades shown in the ID dendrogram and 3 cluster clades shown in the MIC dendrogram. No dominant cluster or cluster combinations were found.

## Discussion

When normally functioning, the lower urinary tract system effectively washes the urethra and bladder with each urination. This prevents the entry of external bacteria or if the bacteria is already inside the bladder, it will be flushed out. Bladder epithelium is also coated with glycosaminoglycan mucin to prevent bacterial adhesion and biofilm formation. Urinary catheter bypasses these defenses, thus introducing bacteria directly from the environment, skin, or gastrointestinal tract into the bladder. Biofilm formation during urinary catheter installation is the most important contributing factor. Crystalline and biofilm formation can obstruct catheter lumen reducing urinary flow. Urease-producing (e.g. *Klebsiella pneumoniae, Proteus spp*.) and mucin-producing bacteria (e.g. *Pseudomonas aeruginosa, Escherichia coli*) as biofilm-producing microorganisms can be prevented universally by implementing catheter-associated urinary tract infection bundles or for high-end use, using antibiotic-impregnated catheter.^15,21^ Hypoxic conditions in COVID-19 patients also permits more biofilm formations.^10,11^ Healthcare-associated infections (HAIs) were also associated with the use of tocilizumab, steroids, hydroxychloroquine, and acute kidney injury requiring hemodialysis.^22^

In this study, more females suffer from CAUTI than males, despite more males being admitted in severe and critical conditions. This is possibly caused by an obvious anatomical factor. The number of CAUTI associated isolates increases with the increase of admitted COVID-19 patients and healthcare worker burdens. Especially in December 2020 and January 2021 in which the first wave of COVID-19 in Indonesia reached its peak.^3^ Thirteen patients died in association with urosepsis caused by CAUTI. This contributes to total COVID-19 deaths, not directly caused by the COVID-19 mechanism, but caused by secondary bacterial infections that are contributed by hospital-associated infections.

Hierarchical clustering is based on high internal homogeneity and high external heterogeneity. If characteristics of phenotype might be triggered or suppressed due to biological, chemical, or physical stressors experienced by the bacteria. Another research showed a dominant cluster when using the same hierarchical clustering analysis towards ventilator-associated pneumonia isolates. This indicates that the isolates have the same phenotypic features that might indicate similar previous stressors. It can be assumed that one cluster of bacteria has undergone a similar stressor that elicits the same phenotype, especially when the MIC is grouped into the same cluster. The same phenotypes also indicate that the same clone was transmitted among the ventilator-associated pneumonia patients. ^16^ This is also supported by the basis of clustering in the dendrogram which explains the outcome of ESBLs and the MDRs that fall into the same criteria as proposed by CLSI.^18,23^ Authors emphasize that this grouping nomenclature and cluster category only applies to this research. A different database can result in different groupings even though the same method is used. This is because of the algorithm of clustering that groups together variables according to their similarities.

The findings in this study showed no dominant cluster, therefore, the stressors experienced by the isolates are different and might be carried from the outside, that is, by the patients. Long catheter-days (9.28 ± 1.37; X □ ± SD) might picture the normal phase of biofilm formation inside the urinary tract after a week of catheter installation. This research is the first to analyze bacterial phenotypic features using hierarchical clustering for CAUTI. This simple method can be applied to empower decision-making and managerial intervention to prevent HAIs, especially CAUTI..

Even though the numbers of the samples are adequate to analyze clusters The limitations of this study are:

## Conclusion

CAUTI associated *E*.*coli* is divided into 9 clusters that did not differ significantly. This indicates no cluster dominates the isolates, thus *E*.*coli* associated CAUTI is less likely caused by hospital transmission of microorganisms but normal flora carried by the admitted patient. This method can be applied to trace common sources of bacterial outbreaks or be used as proof for isolated tracing by phenotypic similarity, especially in limited-resource settings. The use of this method can set a reference for managerial interventions, thus preventing HAI.

## Data Availability

All data produced in the present work are contained in the manuscript

## Acknowledgements

We thank Dr. Soetomo Academic Hospital for providing all necessary support in this research.

## Author Contributions

DE designed the study. DE, PDE designed the methodology. DE performed the experiments. PDE and NMM supervised the experiments. DE, PDE and NMM analysed the data. DE, PDE, and NMM wrote the paper with input from all authors.

